# Effects of SARS-CoV-2 Infection on Attention, Memory, and Sensorimotor Performance

**DOI:** 10.1101/2022.09.22.22280222

**Authors:** Erin E. O’Connor, Nikita Rednam, Rory O’Brien, Shea O’Brien, Peter Rock, Andrea Levine, Thomas A. Zeffiro

**Author notes:** **Corresponding author:** Erin O’Connor, MD, Address: University of Maryland School of Medicine, Department of Diagnostic Radiology & Nuclear Medicine, 22 S. Greene Street, Baltimore, MD 21201. These authors contributed equally to this work and share first authorship (Erin E. O’Connor, Thomas A. Zeffiro). **Study funding:** University of Maryland Baltimore, Institute for Clinical & Translational Research and the National Center for Advancing Translational Sciences (NCATS) CTSA grant number 1UL1TR003098. **Disclosures:**. Dr. O’Connor reports no disclosures. Ms. Rednam reports no disclosures. Mr. O’Brien reports no disclosures. Ms. O’Brien reports no disclosure. Dr. Rock reports no disclosures. Dr. Levine reports no disclosures. Dr. Zeffiro reports no disclosures.

## Abstract

**Background:** Recovery after SARS-CoV-2 infection is extremely variable, with some individuals recovering quickly, and others experiencing persistent long-term symptoms or developing new symptoms after the acute phase of infection, including fatigue, poor concentration, impaired attention, or memory deficits. Many existing studies reporting cognitive deficits associated with SARS-CoV-2 infection are limited by the exclusive use of self-reported measures or a lack of adequate comparison groups.

**Methods:** Forty-five participants, ages 18-70, (11 Long-COVID, 14 COVID, and 20 No-COVID) underwent behavioral testing with the NIH Toolbox Neuro-Quality of Life survey and selected psychometric tests, including a flanker interference task and the d2 Test of Attention.

**Results:** We found greater self-reported anxiety, apathy, fatigue, emotional dyscontrol, sleep disturbance and cognitive dysfunction in COVID compared No-COVID groups. After categorizing COVID patients according to self-reported concentration problems, we observed declining performance patterns in multiple attention measures across No-COVID controls, COVID and Long-COVID groups. COVID participants, compared to No-COVID controls, exhibited worse performance on NIH Toolbox assessments, including the Eriksen Flanker, Nine-Hole Pegboard and Auditory Verbal Learning tests.

**Conclusion:** This study provides convergent evidence that previous SARS-CoV-2 infection is associated with impairments in sustained attention, processing speed, self-reported fatigue and concentration. The finding that some patients have cognitive and visuomotor dysfunction in the absence of self-reported problems suggests that SARS-CoV-2 infection can have unexpected and persistent subclinical consequences.

## Introduction

Recovery after SARS-CoV-2 infection is extremely variable. While some individuals recover quickly, many others experience persistent long-term symptoms or develop new symptoms after the acute phase of infection. Persistent or new signs or symptoms after the acute phase of SARS-CoV-2 infection is known as “Long-COVID”, or post-acute sequelae associated with SARS-CoV-2 infection (PASC). These signs and symptoms include fatigue, dyspnea, chest pain, persistent loss of taste and/or smell, cognitive changes, arthralgias, and decreased quality of life, reflecting the multiple organ systems affected by SARS-CoV-2 infection [1, 2]. Persisting complaints are reported to occur in 20-25% of patients after SARS-CoV-2 infection [3]. Common neuropsychiatric symptoms after the acute phase of infection include fatigue, poor concentration, impaired attention, memory deficits and mood disturbance, suggesting CNS effects [2, 4-11]. These sequelae can adversely affect quality of life [2, 9, 11, 12].

Many existing studies of Long-COVID, however, are limited by the use of self-reported outcomes, selection bias or a lack of adequate control groups. Further, some studies that do incorporate control groups also report these same symptoms in controls, albeit at lower incidence [10, 13]. It is not yet clear whether the presence of self-reported cognitive complaints, such as “brain fog” or concentration problems, accurately identifies individuals with SARS-CoV-2 infection related brain effects. It is possible that psychometric testing could reveal subclinical dysfunction that would be relevant to the problem of understanding the neuropathophysiology of SARS-CoV-2 infection.

In this study we administered quality of life questionnaires and conducted neuropsychological testing to evaluate signs and symptoms and quantify the cognitive and sensorimotor deficits that may occur after SARS-CoV-2 infection. We adjusted for confounding influences such as mood disturbance, comorbidities, and acute infection severity.

## Materials and Methods

### Participants

We recruited 45 participants, ages 18-70, (11 Long-COVID, 14 COVID, and 20 No-COVID) from a post-COVID-19 clinic, the University of Maryland COVID-19 Biorepository, and advertisements. All COVID-19 participants had a positive PCR test and greater than 4 weeks had passed since their initial date of infection. Participants with persistent symptoms were predominantly referred from the post-acute COVID-19 clinic where a clinician with Long-COVID expertise had interviewed and examined them. They were all greater than 16 weeks since initial infection. We interviewed No-COVID control participants regarding potential exposures, absence of cognitive complaints and safety precautions taken during the pandemic before assignment to minimize the possibility of prior asymptomatic SARS-CoV-2 infection. To increase generalizability of our results, use of alcohol, marijuana and nicotine were permitted. Marijuana use was not permitted on the day of, or day prior to, participation. For other substances, including cocaine, methamphetamine, heroin, phencyclidine, and controlled medications, participants were excluded if they had substance use disorder or any regular use in the past 90 days. We screened for these substances on the day of testing with urine drug screens.

Other exclusion criteria included pregnancy, claustrophobia, contraindications to MRI, neurologic disorders unrelated to SARS-CoV-2 infection including seizure disorders, closed head injuries with loss of consciousness greater than 15 minutes, CNS neoplasm, or history of stroke. We also excluded candidates with unstable cardiac or pulmonary disease, and incarcerated prisoners. All participants had a negative rapid antigen test for acute COVID-19 infection on the day of participation.

Study procedures were approved by the Institutional Review Board at University of Maryland School of Medicine and all participants provided written informed consent.

### Questionnaires

All participants answered an online questionnaire to provide demographic information and characterization of their COVID-19 infection illness. Participants answered eight items regarding their dates of infection, whether they had been hospitalized, treatments administered, and vaccination status.

### Behavioral assessments

The following cognitive tests were administered in person via the NIH Toolbox:

▪ The **Neuro-Quality of Life** questions (Neuro-QOL) included self-report measures of health-related quality of life for individuals affected by neurological disorders [14].We included Anxiety, Depression, Apathy, Fatigue, Sleep, Executive Function, and Cognition Domains.
▪ **Nine-Hole Pegboard Dexterity** measured visuomotor coordination in both dominant and non-dominant hands, assessed by placing pegs in a 9-hole pegboard in a timed manner [15, 16].
▪ **Auditory Verbal Learning Test** measured memory for of a list of 15 unrelated words, followed by attempted recall over 3 trials.
▪ **Dimensional Change Card Sort** used color and shape matching to assess cognitive flexibility [17].
▪ The **Flanker Inhibitory Control and Attention Task** measured ability to sustain attention and discern between concordant and discordant cues when presented with flanking stimuli [17]
▪ The **Grip Strength Test** measured individual hand strength in both dominant and non-dominant hands while squeezing a dynamometer [16].
▪ The **List Sorting Working Memory Task** measured working memory when participants were presented with a sequence of pictures with verbal descriptions in order [18].
▪ The **Oral Reading Recognition** measured single word reading, requiring pronunciation of words administered using an adaptive format [19].
▪ The **Picture Vocabulary Test** measured vocabulary when participants listen to a word and selected a picture that best matched the word’s meaning [19].
▪ The **Oral Symbol Digit Test** measured processing speed when participants match symbols to numbers using a legend. Scores were calculated by items accurately completed over 120 seconds [20, 21].
▪ The **Pattern Comparison Test** measured processing speed by asking participants to distinguish two given visuals as same or different [20].
▪ The **Picture Sequence Memory Test** measured episodic memory when participants listened to and watched a sequence of pictures and events, then putting the pictures in the sequence in which they were shown [22]
▪ The **Odor Identification Task** assessed olfactory function using scratch and sniff test cards for matching an odor with its corresponding picture [23].
▪ **The Regional Taste Intensity Test** measured a participant’s sensitivity to taste by tasting mixtures and ranking their intensity on a scale [24].

### The d2 Test of Attention

Patients commonly report “brain fog” with poor concentration in the subacute and chronic phases after SARS-CoV-2 infection, sometimes after mild infection [25]. Poor concentration can be operationalized as attention dysfunction, and we therefore had participants undergo formal psychometric testing. The d2 Test of Attention measures attention and concentration processes by asking participants to differentiate visually similar stimuli in a timed manner [26]. Participants select the target characters (a “d” with a total of two dashes placed above and/or below) with a pencil amongst non-target characters (a “d” with more or less than two dashes, and “p” characters with any number of dashes), in 14 consecutive 20 second trials [26]. For each participant, we computed the total number of items processed (TN), omission errors/d2s omitted (E_1_), commission errors/irrelevant letters crossed out (E_2_), sum of omission and commission errors € and the percentage of errors (E%). The number of correct items (TNE) was computed from TN and E. We computed concentration performance (CP) by subtracting E_2_ from the sum of correctly crossed out relevant items.

### Statistical analysis

Statistical analysis was done using R 2.41 [27]. Effects were modeled using multiple regression, with group, age, sex, and education used as fixed effects and Neuro-QoL reports and cognitive and sensorimotor measures as outcomes. Depression was included in some of the cognitive outcome models. Model results included standardized parameter estimates and their associated confidence intervals.

A Hotelling’s two sample T2-multivariate test was used to examine differences in the Neuro-QoL response profiles.

## Results

### Participants

We saw no evidence that participants differed in age, sex, or education (Table 1). Eleven participants had persistent cognitive symptoms after the acute phase of SARS-CoV-2 infection and were classified as Long-COVID. Those without complaints of concentration problems or mental fatigue were included in the COVID group. One participant had SARS-CoV-2 infection twice but was asymptomatic after acute infection in both instances. No participants failed the drug screen on the day of the exam.

**Table 1.**
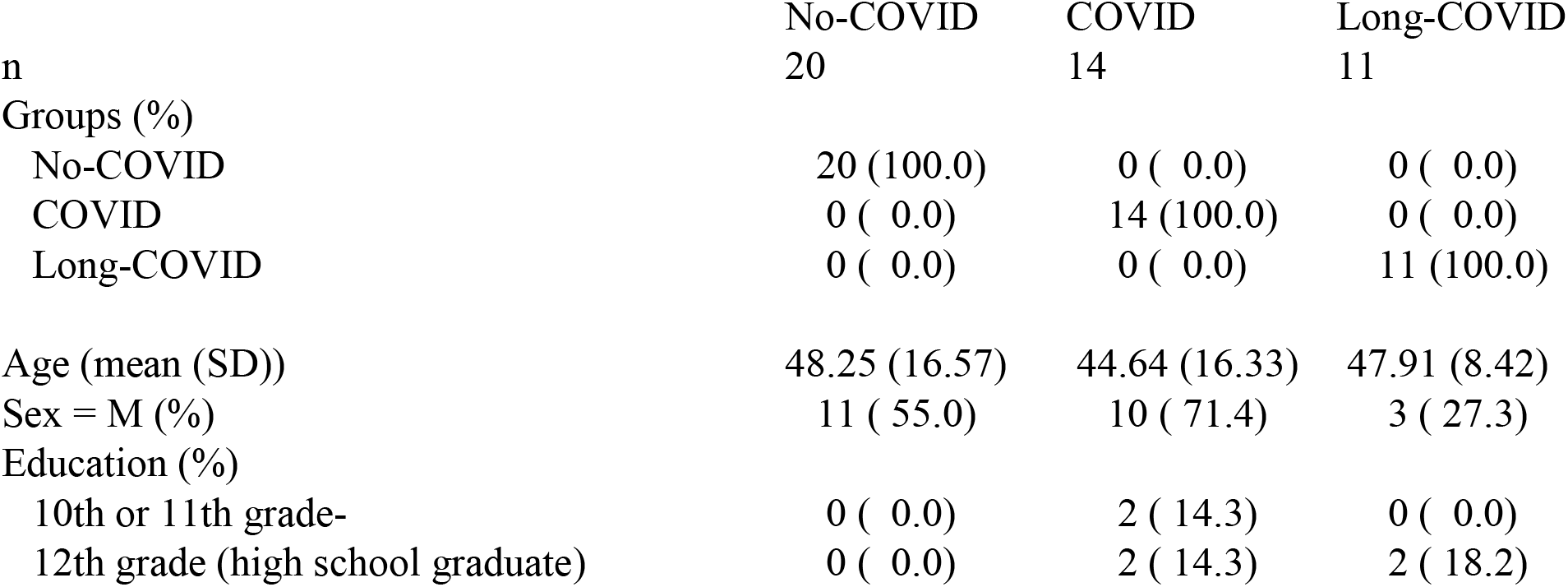
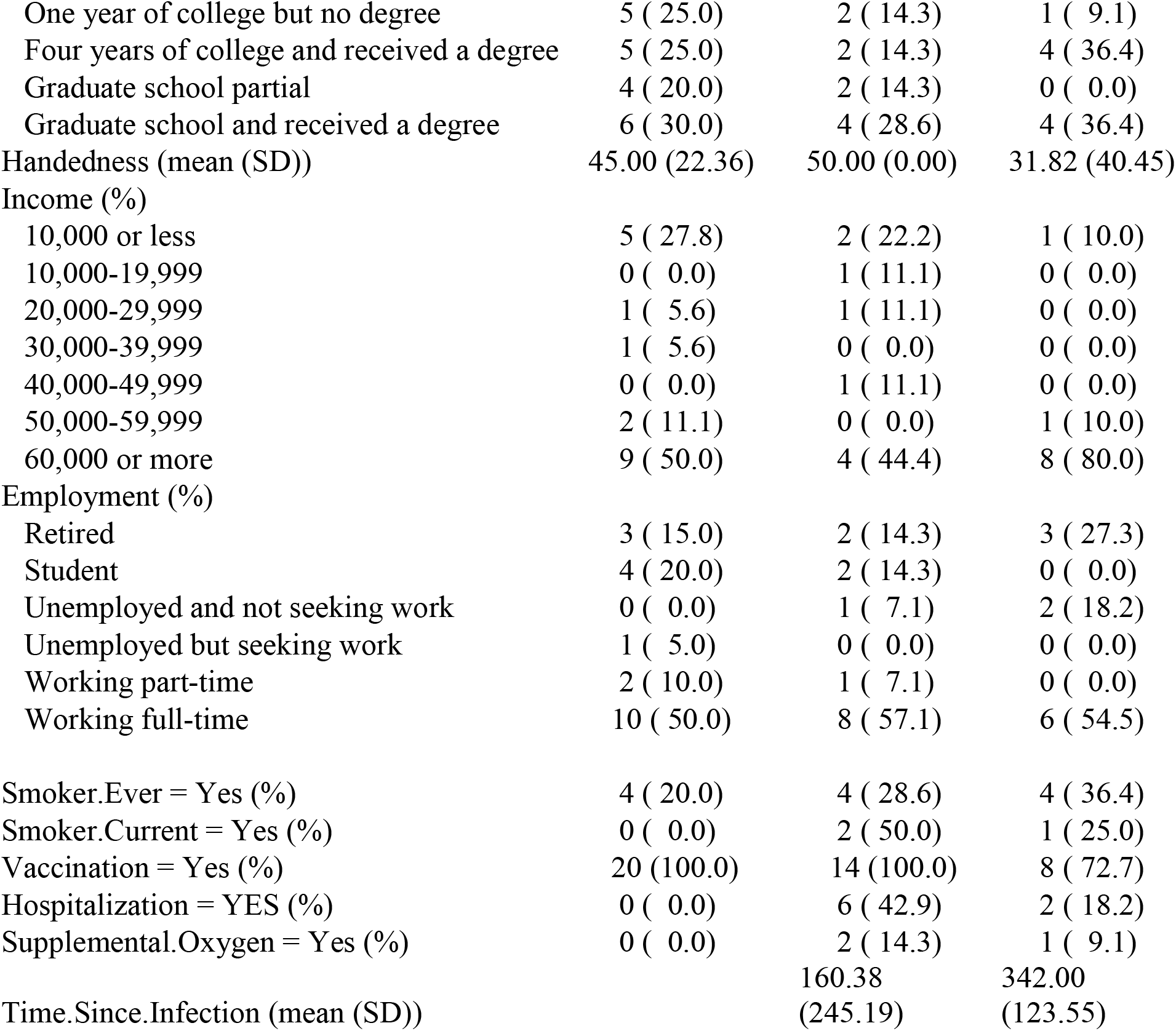
Participant Characteristics

### Behavioral Assessments

We next examined the magnitude of self-reported symptoms in the 3 groups. The NIH Toolbox Neuro-QoL scores revealed differences among No-COVID, COVID and Long-COVID participants. The Long-COVID group exhibited greater anxiety, apathy, fatigue, sleep disturbance, and emotional dyscontrol than the No-COVID group (Figure 1). They also experienced diminished cognitive function. The Long-COVID vs No-COVID comparisons were all statistically significant except for depression.

**Figure 1.**
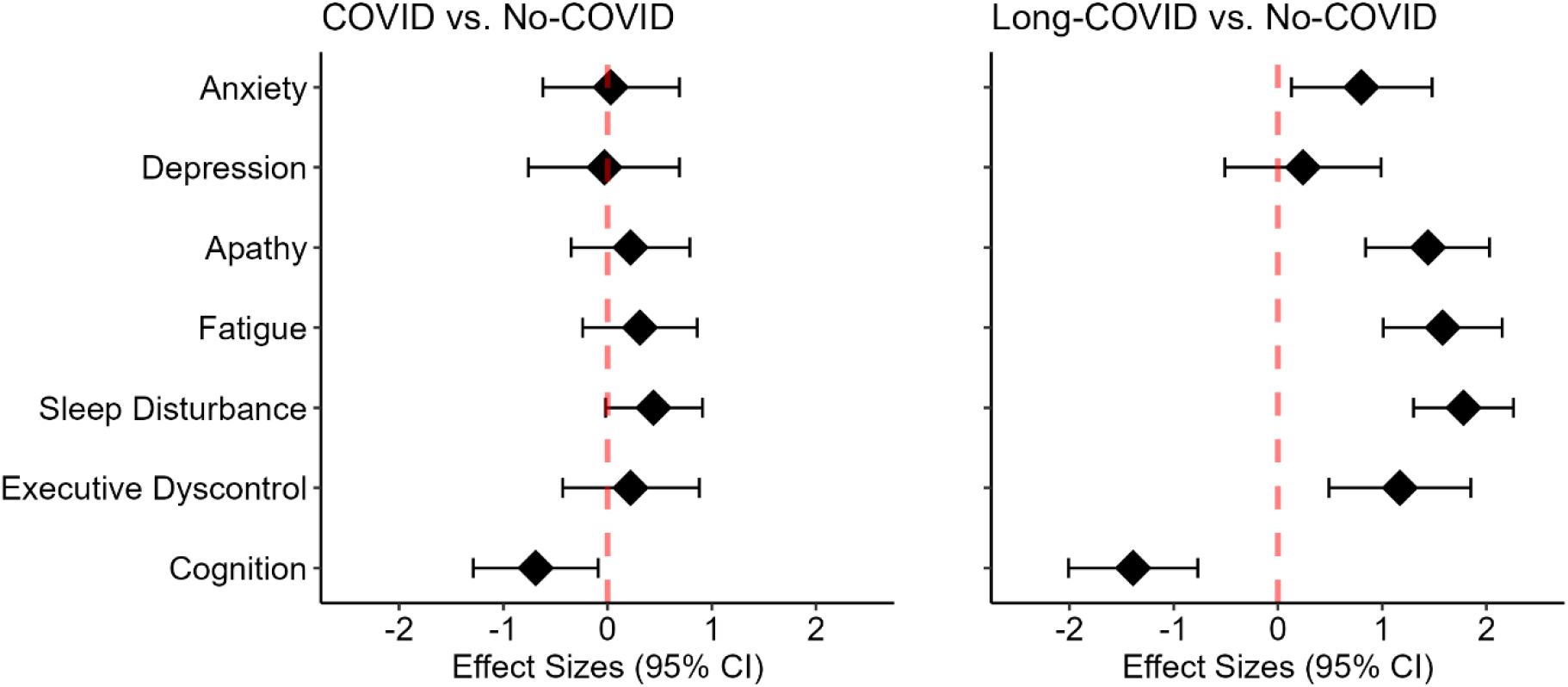
NIH Toolbox Neuro-QOL measures revealed greater anxiety, apathy, fatigue, emotional dyscontrol, sleep and cognitive disturbances in the Long-COVID group than the No-COVID controls.

A multivariate Hotelling’s two sample T2-test provided evidence for a difference in the Long-COVD vs COVID profiles: T.2 = 6.48, df1 = 7, df2 = 18, p-value <0.01; the alternative hypothesis was that the true profile difference was not equal to (0,0,0,0,0,0,0). *d*2 Test of Attention. Using age, sex, and education as covariates in our models, the COVID groups exhibited impairments in both attention and processing speed, evidenced by lower numbers of Correct Items (Long-COVID vs. No-COVID, *β* = -0.82, CI = [-1.53 – -0.10], COVID vs. No-COVID, *β* = -0.92, CI =[ -1.61 – -0.22]) (Figure 2A); poorer Concentration Performance, a measure of sustained attention (Long-COVID vs. No-COVID, *β* = -0.69, CI = [-1.42 – 0.03], COVID vs. No-COVID, *β* = -0.73, CI =[-1.44 – -0.03]) (Figure 2B); and lower total numbers of Items Processed, a measure of processing speed (Long-COVID vs. No-COVID, *β* = -0.86, CI = [-1.58 – -0.14], COVID vs. No-COVID, *β* = -0.96, CI = [ -1.66 – -0.26]) (Figure 2C). No differences were seen directly comparing the Long-COVID and COVID groups. The No-COVID group had the largest number of correct items, followed by the COVID group and the Long-COVID groups.

**Figure 2.**
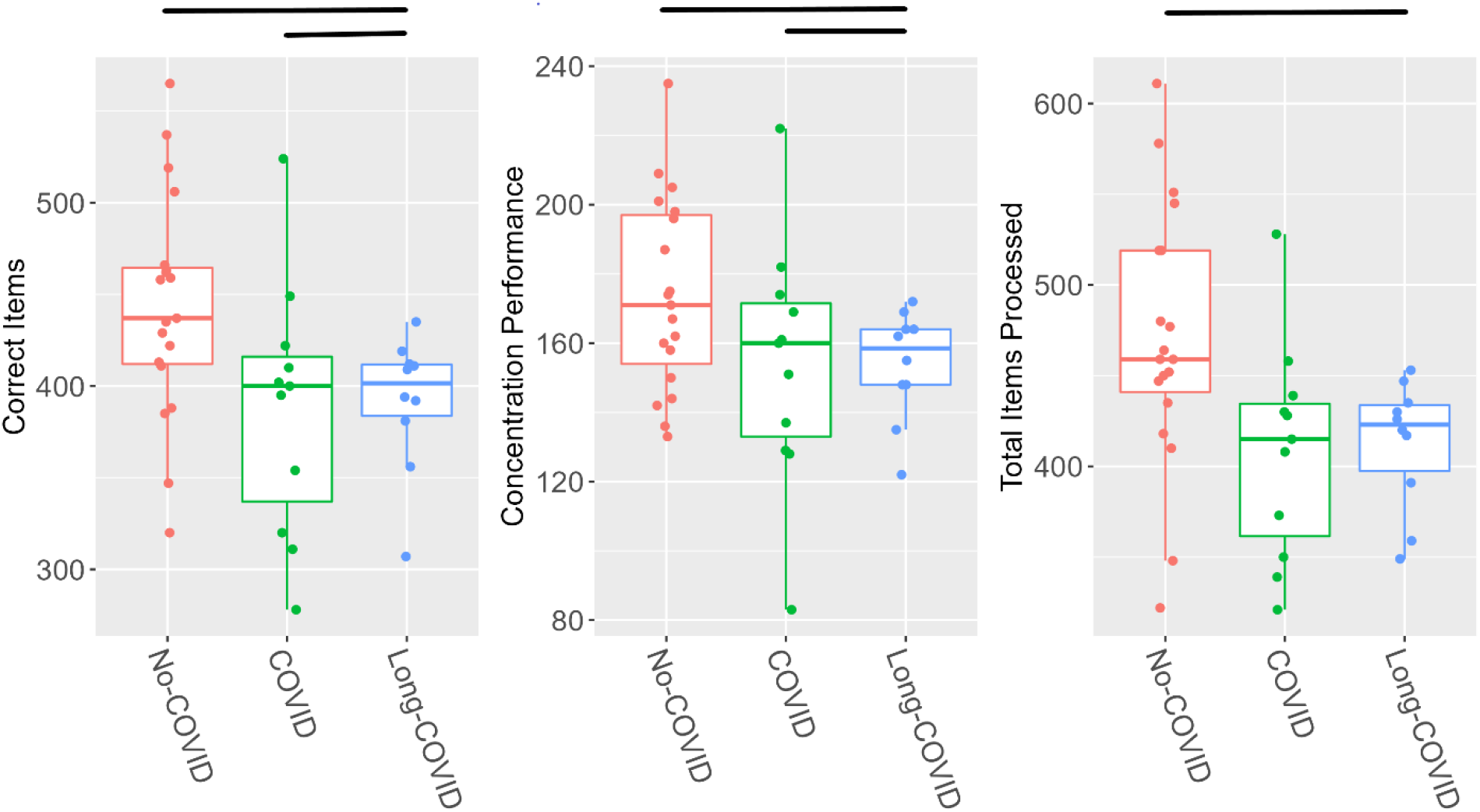
The COVID group exhibited poorer performance than the No-COVID controls in (a) number of Correct Items, (B) Concentration Performance and (C) Items Processed in the d2 Attention Test. Horizontal bars show statistical between-group differences.

### NIH Toolbox Cognitive Measures

Eriksen Flanker Task. Consistent with the attention deficits observed using the d2 Test of Attention, both patients groups took longer to complete the Eriksen Flanker Task (Long-COVID vs. No-COVID, *β* =0.46, CI = [-0.17 – 1.10], COVID vs. No-COVID, *β* =0.73, CI = [0.12 – 1.33]). They also had lower Eriksen Flanker computed scores (Long-COVID vs. No-COVID, *β* =-0.50, CI = [-1.19 – 0.18], COVID vs. No-COVID, *β* = -0.94, CI = [-1.61 – -0.26]) (Figure 3). The computed scores combine information about both speed and accuracy. We again used age, sex, and education as covariates in our models. We failed to observe evidence that the Long-COVID and COVID groups differed statistically. (Figure 3).

**Figure 3.**
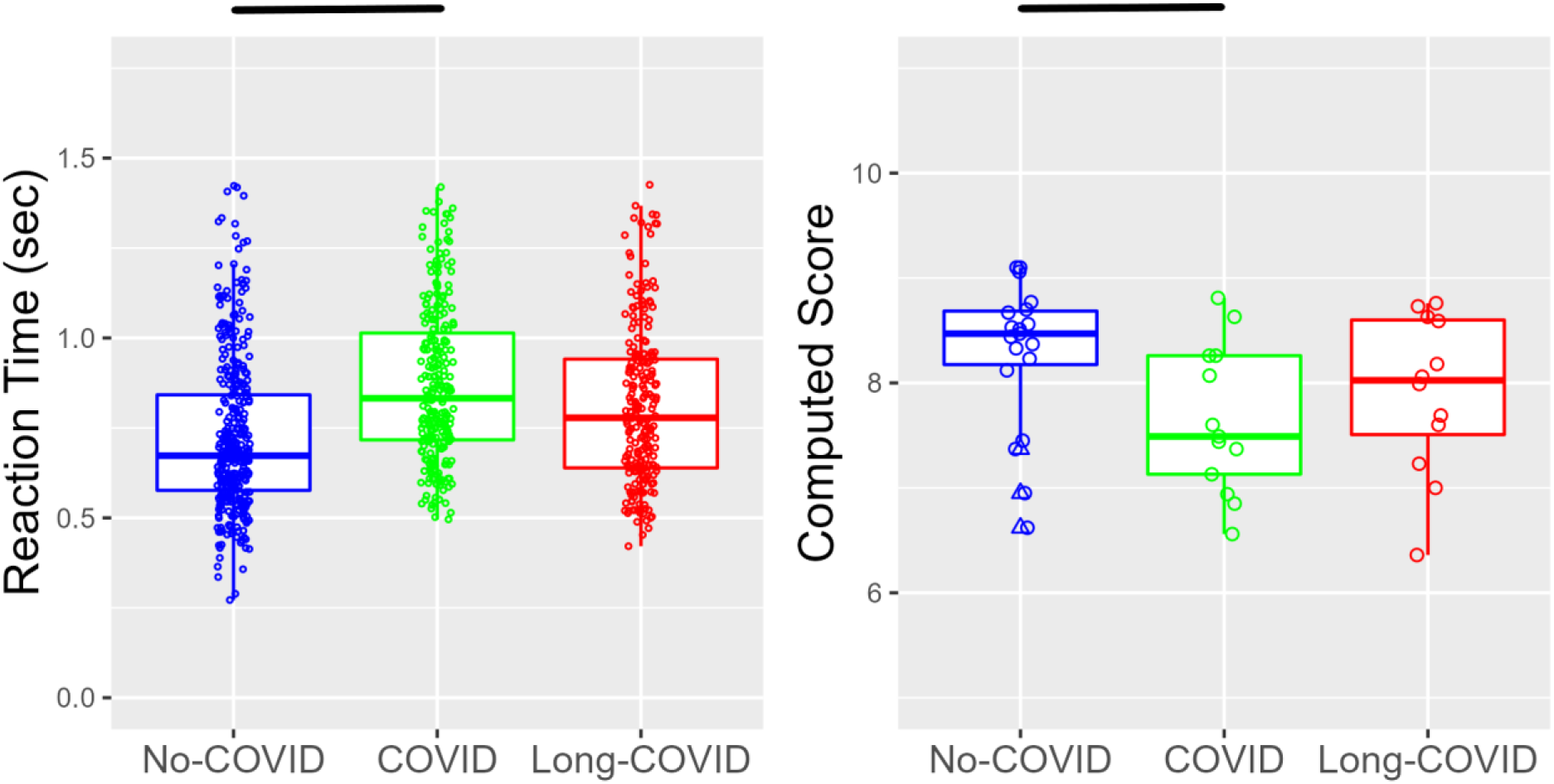
The COVID and Long-COVID group had slower reaction times (A) and had lower computed scores (B) on the NIH Toolbox Eriksen Flanker Task. Horizontal bars show statistical between-group differences.

Verbal Learning. We also observed deficits associated with SARS-CoV-2 infection in the Auditory Verbal Learning Test, (Long-COVID vs. No-COVID, *β* = -0.67, CI = [-1.36 – 0.03], COVID vs. No-COVID, *β* = - -0.99, CI = [-1.65 – -0.33]) (Figure 4). Again, we failed to observe evidence that the Long-COVID and COVID groups differed statistically. Including depression in the model did not account for any significant variance.

**Figure 4.**
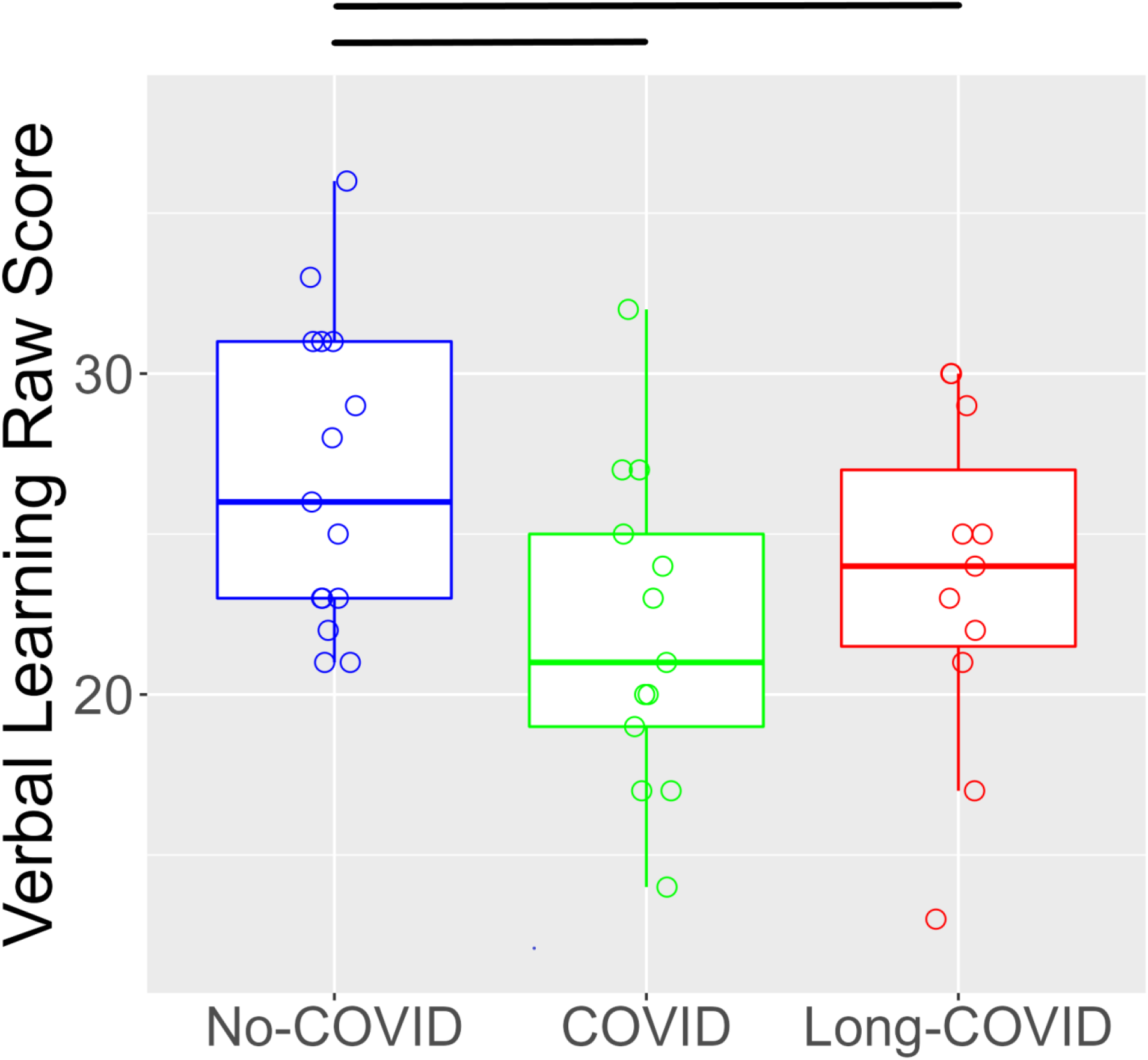
Both Long-COVID and COVID participants performed worse than No-COVID participants on the Auditory Verbal Learning Test, an index of declarative memory. Horizontal bars show statistical between-group differences.

Pegboard Test. Finally, we observed deficits in visuomotor coordination, with both patient groups taking longer to complete the Nine-Hole Pegboard with dominant (Long-COVID vs. No-COVID, *β* = 1.22, CI = [0.56 – 1.89], COVID vs. No-COVID, *β* = 0.50, CI =[ -0.13 – 1.14]) and non-dominant hands (Long-COVID vs. No-COVID, *β* = 1.02, CI = [0.29 – 1.76], COVID vs. No-COVID, *β* = 0.69, CI =[-0.01 – 1.39]) (Figure 5).

**Figure 5.**
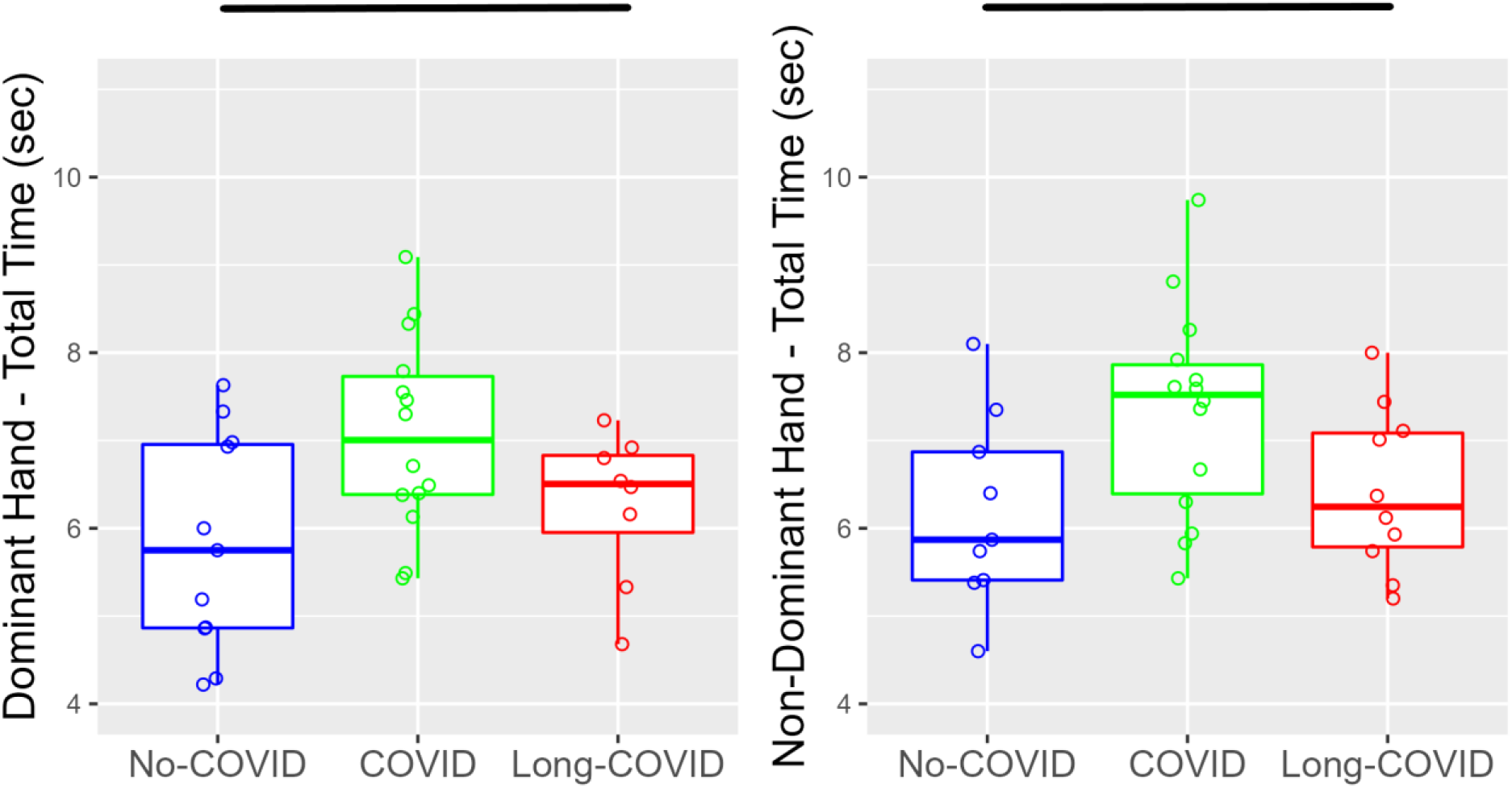
Long-COVID and COVID participants performed worse than No-COVID participants on the Nine-Hole Pegboard with both dominant and non-dominant hands. Horizontal bars show statistical between-group differences.

We did not observe group differences in the other NIH Toolbox test results, including Dimensional Change Card Sort, Grip Strength, List Sorting Working Memory, Oral Reading Recognition, Picture Vocabulary, Oral Symbol Digit, Pattern Comparison, Picture Sequence Memory, Odor Identification and Regional Taste Intensity Tests.

## Discussion

Summary of results. This study examined neurologic quality of life and neuropsychological differences >14 weeks after SARS-CoV-2 infection. While quality of life changes estimated by self-report can be global and substantial following SARS-CoV-2 infection, the degree of impairment across the different neuropsychological domains varies. Our study found convergent evidence for effects of SARS-CoV-2 infection on attention, processing speed, visuomotor coordination and declarative memory. Some COVID participants without self-reported cognitive complaints showed deficits on psychometric testing, suggesting that SARS-CoV-2 infection can have subclinical effects.

We observed convergent evidence for attention dysfunction associated with SARS-CoV-2 infection. The d2 Test of Attention results provide evidence of impairments in sustained attention, selective attention, and processing speed in participants with previous SARS-CoV-2 infection. Another attention test, the Eriksen Flanker Task revealed further evidence of attentional dysfunction related to SARS-CoV-2 infection. The SARS-CoV-2 infected group also performed worse on the Nine-Hole Pegboard Test. While pegboard performance is primarily a measure of visuomotor function, the patients did not exhibit more basic motor abnormalities, as evidenced by the absence of between-group grip strength differences. Attention deficits may also have contributed to poorer performance on Nine-Hole Pegboard [28, 29].

Although other studies have reported cognitive deficits after the acute phase of SARS-CoV-2 infection, few of these studies used quantitative measures for assessment. In a recent meta-analysis that included 43 studies reporting cognitive impairment following resolution of acute COVID-19, only 15 used quantitative measures in their assessment. Amongst the quantitative assessments, many did not have controls and were limited to hospitalized patients [30-42]. Several of the quantitative assessment studies used the MoCA (Montreal Cognitive Assessment) or other screening assessments for cognitive impairment, such as Screen for Cognitive Impairment in Psychiatry (SCIP), Brief Assessment of Cognition in Schizophrenia (BACS), or the Orientation-Memory-Concentration Screening Test, without more in-depth neuropsychological evaluation [30-37, 40-42]. Other studies performed a more in-depth assessment via phone using the Telephone Assessment of Cognitive Status (TICS) or had participants complete testing online without supervision [25, 38, 39, 43]. Morin et al. reported cognitive impairment, measured by the MoCA or a revised d2 Test of Attention score in 38% of patients after hospital discharge for SARS-CoV-2 infection. This study did not provide details regarding the proportion of individuals diagnosed by the MoCA as opposed to the d2 Test of Attention, and performance scores for the various aspects of the d2 were not reported, limiting comparison with our findings [34]. Our findings agree with Zhao et al. who found deficits in episodic memory and sustained attention using unsupervised online psychometric tests in individuals who had asymptomatic to SARS-CoV-2 infection [43]. We also found deficits in selective attention, sustained attention and visuomotor coordination in our cohort. It is possible that infection severity contributed to our findings, given that some of our participants required hospitalization.

One quantitative study found no cognitive impairment in a group of mild to moderate COVID patients compared to controls 4 months after SARS-CoV-2 infection using the Mini Mental Status Exam [44]. It is possible that other ongoing sequelae of SARS-CoV-2 infection were not captured by the outcome measures used.

Our COVID participants included a range of infection severity, with 30.8% requiring hospitalization, but many others only experiencing mild to moderate infection. We found evidence of greater impairments in attention and processing speed related to the severity of the self-reported fatigue and concentration problems in participants with previous SARS-CoV-2 infection. This argues against a categorical effect of SARS-CoV-2 infection. Instead, we believe that infection can result in a spectrum of cognitive differences in all infected individuals, with individuals diagnosed with Long-COVID on the more severe end of this spectrum. Self-reported impairment was not associated with hospitalization. To our knowledge, there is no objective biomarker that predicts the observed variation in cognitive and sensorimotor effects following SARS-CoV-2 infection. This raises the question of whether SARS-CoV-2 affects the brain in all infected individuals, with variation in the degree of brain injury and immune response or whether SARS-CoV-2 infection affects the brain in only a subgroup of infected individuals.

Our study has several strengths. First, we performed in-depth psychometric evaluation to precisely characterize cognitive and visuomotor deficits associated with SARS-CoV-2 infection. Second, we conducted our evaluation in person, rather than by phone or online, ensuring that participants gave adequate effort to complete test items correctly. Third, use of the d2 Test of Attention and the Eriksen Flanker task allowed us to explore different facets of attention including selective and sustained attention, along with visual scanning speed, providing convergent evidence for attention effects. Finally, our patients exhibited a range of infection severity, some with and some without persistent symptoms. Using this approach, we were able to demonstrate a continuous spectrum of cognitive differences related to SARS-CoV-2 infection.

This finding may have particular importance because of the absence of a consensus regarding the definition of Long-COVID.

### Limitations

This study has some limitations. First, our small sample size may have led to a type II error. For this reason, we provided effect sizes and confidence intervals to give estimates of effects to be expected in a larger sample. Second, some of our participants were referred from a post-COVID clinic and were clinically diagnosed with Long-COVID, which may have led to an overestimation of impairment following SARS-CoV-2 infection. Finally, it is possible that some of our No-COVID controls previously had mild and undiagnosed SARS-CoV-2 infection. To overcome this, we selected controls who had been asymptomatic and exercised extreme caution in mitigating the risk of acquiring infection.

## Conclusion

Our findings add to the emerging quantitative evidence for persisting cognitive and visuomotor consequences of SARS-CoV-2 infection, emphasizing the convergent evidence for attention dysfunction. While poor concentration and mental fatigue are major complaints in patients with Long-COVID, little is known about the underlying neuropathophysiological mechanisms [12]. Surprisingly, SARS-CoV-2 infection can have subclinical cognitive consequences. Further studies are needed to determine if brain structural and functional impairment are related to the behavioral complaints and performance deficits seen long after acute SARS-CoV-2 infection.

## Data Availability

All data produced in the present study are available upon reasonable request to the authors

## Author Contributions

The authors confirm contribution to the manuscript as follows: study conception and design: EO, TZ; data collection: EO, NR, PR, AL, RO, SO; analysis and interpretation of results: EO, NR, TZ. All authors reviewed the results and approved the final version of the manuscript.

## Acknowledgments

This study was funded by the University of Maryland Baltimore, Institute for Clinical & Translational Research and the National Center for Advancing Translational Sciences (NCATS) CTSA grant number 1UL1TR003098. The authors would like to acknowledge the study participants for their time and effort that made this study possible. We would also like to acknowledge LaToya Stubbs and Kendra Petrick, research coordinators for the University of Maryland COVID-19 Biorepository.

